## Supplementary for "Patient-specific multi-modal modeling uncovers neurotransmitter receptor involvement in motor and non-motor axes of Parkinson’s disease"

**Supplementary Table S1**: Summary of demographic data for N=71 PD patients.

| **Category** | **PD subjects** |
| --- | --- |
| **Mean MDS-UPDRS Part III score** | 18.8 ± 8.7 |
| **Female patients** | 20 (28.2%) |
| **Mean age (years)** | 59.6 ± 9.8 |
| **Mean education (years)** | 15.5 ± 2.8 |
| **Non-white patients** | 0 |
| **Right handed patients** | 64 (90.1%) |

**Supplementary Table S2:** *Mean and standard deviation of the number of clinical evaluations per subject.*

| **Category** | **Number of evaluations** |
| --- | --- |
| **BJLOT** | 7.62 ± 1.13 |
| **GDS** | 8.27 ± 1.09 |
| **HVLT** | 7.65 ± 1.14 |
| **LNS** | 7.63 ± 1.12 |
| **LXF** | 0.79 ± 0.56 |
| **NP1** | 15.0 ± 1.9 |
| **NP2** | 15.0 ± 1.9 |
| **NP3** | 19.5 ± 4.0 |
| **NP4** | 9.94 ± 3.0 |
| **MoCA** | 7.63 ± 1.12 |
| **SF** | 7.62 ± 1.11 |
| **STAIAD** | 8.28 ± 1.08 |
| **SDM** | 7.66 ± 1.15 |

**Supplementary Table S3:** *Neurotransmitter receptor ligands used to obtain receptor maps.*

| **Neurotransmitter** | **Receptor** | **Ligand** | **Type** |
| --- | --- | --- | --- |
| **Glutamate** | AMPA | [${}^{3}$H]-AMPA | Agonist |
|  | NMDA | [${}^{3}$H]-MK-801 | Antagonist |
|  | Kainate | [${}^{3}$H]-Kainate | Agonist |
| **GABA** | GABA_A_ | [${}^{3}$H]-Muscimol | Agonist |
|  | GABA_B_ | [${}^{3}$H]-CGP 54626 | Antagonist |
|  | GABA_A_-associated benzodiazepine binding site (GABA_A_/BZ) | [${}^{3}$H]-Flumazenil | Antagonist |
| **Acetylcholine** | M_1_ | [${}^{3}$H]-Pirenzepine | Antagonist |
|  | M_2_ | [${}^{3}$H]-Oxotremorine-M | Agonist |
|  | M_3_ | [${}^{3}$H]-4-DAMP | Antagonist |
|  | Nicotinic α_4_β_2_ | [${}^{3}$H]-Epibatidine | Agonist |
| **Noradrenaline** | α_1_ | [${}^{3}$H]-Prazosin | Antagonist |
|  | α_2_ | [${}^{3}$H]-RX 821002 | Antagonist |
| **Serotonin** | 5-HT_1A_ | [${}^{3}$H]-8-OH-DPAT | Agonist |
|  | 5-HT_2_ | [${}^{3}$H]-Ketanserin | Antagonist |
| **Dopamine** | D_1_ | [${}^{3}$H]-SCH 23390 | Antagonist |

**Supplementary Table S4**: *Brain regions with receptor data, and the corresponding atlas used to extract the ROI map. Note that regions are defined by cytoarchitecture, and thus do not correspond perfectly with functional regions.*

| **Lobe** | **Anatomical subdivision** | **Jülich area** | **Region name** | **Atlas source** |
| --- | --- | --- | --- | --- |
| **Occipital lobe** | Visual cortex | hOc1 | Brodmann’s area 17 / V1 | Jülich |
|  |  | hOc2 | Brodmann’s area 18 / V2 | Jülich |
|  |  | hOc4d | V4 | Jülich |
|  |  | hOc3a | V3a | Jülich |
|  |  | hOc3d | V3d | Jülich |
|  |  | hOc3v | V3v | Jülich |
|  |  | hOc4v | V4 | Jülich |
|  | Extrastriate cortex | FG1 | Part of Brodmann area 19 | Jülich |
|  |  | FG2 | Part of Brodmann area 19 | Jülich |
| **Parietal lobe** | Somatosensory cortex | 1 | Brodmann’s area 1 | Jülich |
|  |  | 2 | Brodmann’s area 2 | Jülich |
|  |  | 3a | Brodmann’s area 3a | Jülich |
|  |  | 3b | Brodmann’s area 3b | Jülich |
|  | Superior parietal lobule | 5L | Brodmann’s area 5L | Jülich |
|  |  | 5M | Brodmann’s area 5M | Jülich |
|  |  | 7A | Brodmann’s area 7A | Jülich |
|  | Inferior parietal lobule | PGa | Anterior inferior parietal area | Jülich |
|  |  | PGp | Posterior inferior parietal area | Jülich |
|  |  | PFt | Temporal inferior parietal area | Jülich |
|  |  | PFm | Medial inferior parietal area | Jülich |
| **Temporal lobe** | Auditory cortex | Te1 | Temporal area 1 (part of Brodmann’s area 41) | Jülich |
|  |  | Te2 | Temporal area 2 (part of Brodmann’s area 41) | Jülich |
|  | Hippocampus | CA | Cornu ammonis | Jülich |
|  |  | DG | Dentate gyrus | Jülich |
|  | Subiculum | Subiculum | Subiculum | Jülich |
|  | Entorhinal cortex | Ent | Brodmann’s area 28 | Jülich |
|  |  | 20 | Brodmann’s area 20 | Brodmann |
|  |  | 21 | Brodmann’s area 21 | Brodmann |
|  |  | 22 | Brodmann’s area 22 | Brodmann |
|  |  | 36 | Brodmann’s area 36 | Brodmann |
|  |  | 37 | Brodmann’s area 37 | Brodmann |
|  |  | 38 | Brodmann’s area 38 | Brodmann |
| **Frontal lobe** | Agranular premotor cortex | 6 | Brodmann’s area 6 | Jülich |
|  | Primary motor cortex | 4p | Brodmann’s area 4p | Jülich |
|  | Broca’s region | 44 |  | Jülich |
|  |  | 45 |  | Jülich |
|  | Frontopolar cortex | Fp1 | Frontopolar area (part of Brodmann area 10) | Jülich |
|  |  | Fp2 | Frontopolar area (part of Brodmann area 10) | Jülich |
|  | Orbitofrontal cortex | Fo1 | Orbitofrontal area (part of Brodmann area 11) | Jülich |
|  | Lateral prefrontal | 46 | Brodmann’s area 46 | Brodmann |
|  |  | 47 | Brodmann’s area 47 | Brodmann |
|  |  | 8 | Brodmann’s area 8 | Brodmann |
|  |  | 9 | Brodmann’s area 9 | Brodmann |
| **Cingulate regions (multiple lobes)** | Anterior cingulate | p24ab | Pregenual cingulate areas p24a & p24b | Jülich |
|  |  | p32 | Pregenual cingulate area p32 | Jülich |
|  | Posterior cingulate | 23 | Brodmann’s area 23 | Brodmann |
|  |  | 31 | Brodmann’s area 31 | Brodmann |
| **Basal ganglia** | Striatum | Putamen | Putamen | AAL |
|  |  | Caudate | Caudate nucleus | AAL |
|  | Pallidum | Globus pallidus | Globus pallidus | DISTAL |
|  | Subthalamic nucleus | STN | Subthalamic nucleus | DISTAL |
| **Forebrain** | Thalamus | Thalamus (anterior) | Thalamus (anterior) | AAL |
|  |  | Thalamus (medial) | Thalamus (medial) | AAL |
|  |  | Thalamus (lateral) | Thalamus (lateral) | AAL |

**Supplementary Table S5:** *Biological parameters most correlated with clinical symptoms in PD via PC1, and the percentage of clinical score covariance explained.*

| **Neuroimaging Modality** | **Model Parameter** | **Receptor Type** | **Explained Variance** |
| --- | --- | --- | --- |
| GM | AMPA x fALLF | Glutamatergic | 0.12% |
|  | GABA_B_ | GABAergic | 0.18% |
|  | α_4_β_2_ x GM | Cholinergic | 0.31% |
|  | M_1_ x fALLF | Cholinergic | 0.10% |
|  | M_2_ x fALLF | Cholinergic | 0.31% |
|  | α_4_β_2_ x fALLF | Cholinergic | 0.10% |
|  | M_3_ x FA | Cholinergic | 0.16% |
|  | M_1_ x t1/t2 | Cholinergic | 0.12% |
|  | M_1_ | Cholinergic | 0.10% |
|  | α_2_ x fALLF | Adrenergic | 0.42% |
|  | 5HT_1A_ x SPECT | Serotonergic | 0.29% |
|  | D_1_ x fALLF | Dopaminergic | 0.29% |
|  | GM | Non-Receptor | 0.14% |
|  | SPECT | Non-Receptor | 0.20% |
| fALFF | Kainate x SPECT | Glutamatergic | 0.10% |
|  | NMDA x FA | Glutamatergic | 0.33% |
|  | Kainate x t1/t2 | Glutamatergic | 0.24% |
|  | Bz site x GM | GABAergic | 0.31% |
|  | GABA_B_ x fALLF | GABAergic | 0.29% |
|  | GABA_A_ x FA | GABAergic | 0.27% |
|  | M_2_ x FA | Cholinergic | 0.13% |
|  | M_1_ | Cholinergic | 0.18% |
|  | M_2_ | Cholinergic | 0.21% |
|  | 5HT_1A_ x t1/t2 | Serotonergic | 0.08% |
|  | D_1_ x GM | Dopaminergic | 0.19% |
|  | GM | Non-Receptor | 0.16% |
| SPECT | Kainate x FA | Glutamatergic | 0.15% |
|  | α_1_ x FA | Adrenergic | 0.12% |
|  | 5HT_2_ x GM | Serotonergic | 0.18% |
|  | 5HT_1A_ x MD | Serotonergic | 0.25% |
|  | 5HT_2_ x t1/t2 | Serotonergic | 0.23% |
|  | 5HT_2_ | Serotonergic | 0.21% |
|  | D_1_ x FA | Dopaminergic | 0.14% |
|  | D_1_ | Dopaminergic | 0.21% |
|  | FA | Non-Receptor | 0.31% |
|  | MD | Non-Receptor | 0.19% |
| FA | Kainate x GM | Glutamatergic | 0.22% |
|  | AMPA x t1/t2 | Glutamatergic | 0.13% |
|  | Kainate x t1/t2 | Glutamatergic | 0.18% |
|  | AMPA | Glutamatergic | 0.36% |
|  | Kainate | Glutamatergic | 0.28% |
|  | GABA_A_ x MD | GABAergic | 0.20% |
|  | Bz site x t1/t2 | GABAergic | 0.20% |
|  | GABA_B_ x t1/t2 | GABAergic | 0.30% |
|  | GABA_A_ | GABAergic | 0.67% |
|  | M_1_ x fALLF | Cholinergic | 0.15% |
|  | M_3_ x MD | Cholinergic | 0.48% |
|  | α_4_β_2_ x t/1t2 | Cholinergic | 0.19% |
|  | α_2_ x GM | Adrenergic | 0.25% |
|  | α_2_ x MD | Adrenergic | 0.23% |
|  | α_1_ x t1/t2 | Adrenergic | 0.16% |
|  | α_1_ | Adrenergic | 0.13% |
|  | 5HT_1A_ x SPECT | Serotonergic | 0.08% |
|  | 5HT_2_ x SPECT | Serotonergic | 0.18% |
|  | 5HT_2_ x MD | Serotonergic | 0.17% |
|  | 5HT_2_ x t1/t2 | Serotonergic | 0.12% |
|  | GM | Non-Receptor | 0.28% |
|  | SPECT | Non-Receptor | 0.12% |
|  | MD | Non-Receptor | 0.21% |
|  | t1/t2 | Non-Receptor | 0.23% |
|  | spreading | Non-Receptor | 0.17% |
| MD | AMPA x fALLF | Glutamatergic | 0.23% |
|  | Kainate x fALLF | Glutamatergic | 0.37% |
|  | Kainate x FA | Glutamatergic | 0.20% |
|  | NMDA x MD | Glutamatergic | 0.25% |
|  | Kainate x MD | Glutamatergic | 0.38% |
|  | GABA_A_ x GM | GABAergic | 0.41% |
|  | Bz site x fALLF | GABAergic | 0.31% |
|  | GABA_B_ x MD | GABAergic | 0.11% |
|  | GABA_A_ | GABAergic | 0.35% |
|  | Bz site | GABAergic | 0.50% |
|  | M_1_ x MD | Cholinergic | 0.18% |
|  | M_1_ | Cholinergic | 0.21% |
|  | M_2_ | Cholinergic | 0.43% |
|  | M_3_ | Cholinergic | 0.67% |
|  | α_1_ x FA | Adrenergic | 0.10% |
|  | α_2_ | Adrenergic | 0.13% |
|  | 5HT_1A_ x GM | Serotonergic | 0.12% |
|  | 5HT_2_ x fALLF | Serotonergic | 0.54% |
|  | 5HT_1A_ x FA | Serotonergic | 0.22% |
|  | 5HT_2_ | Serotonergic | 0.45% |
|  | D_1_ x FA | Dopaminergic | 0.15% |
|  | fALLF | Non-Receptor | 0.29% |
|  | FA | Non-Receptor | 0.24% |
| t1/t2 | AMPA x FA | Glutamatergic | 0.21% |
|  | NMDA x FA | Glutamatergic | 0.55% |
|  | NMDA | Glutamatergic | 0.38% |
|  | Kainate | Glutamatergic | 0.16% |
|  | Bz site x GM | GABAergic | 0.31% |
|  | GABA_A_ x FA | GABAergic | 0.62% |
|  | Bz site x FA | GABAergic | 0.27% |
|  | Bz site x MD | GABAergic | 0.35% |
|  | Bz site x t1/t2 | GABAergic | 0.13% |
|  | GABA_B_ x t1/t2 | GABAergic | 0.20% |
|  | M_1_ x SPECT | Cholinergic | 0.21% |
|  | M_1_ x FA | Cholinergic | 0.15% |
|  | M_2_ x FA | Cholinergic | 0.08% |
|  | M_1_ x t1/t2 | Cholinergic | 0.13% |
|  | M_3_ x t1/t2 | Cholinergic | 0.18% |
|  | α_2_ x fALLF | Adrenergic | 0.28% |
|  | α_2_ | Adrenergic | 0.21% |
|  | 5HT_2_ x GM | Serotonergic | 0.52% |
|  | 5HT_2_ x SPECT | Serotonergic | 0.14% |
|  | D_1_ x FA | Dopaminergic | 0.19% |
|  | offset | Non-Receptor | 0.25% |
|  | FA | Non-Receptor | 0.19% |

**Supplementary Table S6:** *Biological parameters most correlated with clinical symptoms in PD via PC2, and the percentage of clinical score covariance explained.*

| **Neuroimaging Modality** | **Model Parameter** | **Receptor Type** | **Explained Variance** |
| --- | --- | --- | --- |
| GM | NMDA x GM | Glutamatergic | 0.06% |
|  | NMDA x SPECT | Glutamatergic | 0.10% |
|  | NMDA x MD | Glutamatergic | 0.05% |
|  | Kainate x MD | Glutamatergic | 0.04% |
|  | AMPA | Glutamatergic | 0.08% |
|  | NMDA | Glutamatergic | 0.04% |
|  | GABA_A_ x GM | GABAergic | 0.04% |
|  | GABA_A_ x fALLF | GABAergic | 0.09% |
|  | GABA_A_ x FA | GABAergic | 0.05% |
|  | GABA_A_ x MD | GABAergic | 0.05% |
|  | M_1_ x fALLF | Cholinergic | 0.07% |
|  | M_1_ x MD | Cholinergic | 0.19% |
|  | M_3_ | Cholinergic | 0.08% |
|  | α_4_β_2_ | Cholinergic | 0.08% |
|  | α_2_ x SPECT | Adrenergic | 0.11% |
|  | α_1_ | Adrenergic | 0.07% |
|  | 5HT_2_ x FA | Serotonergic | 0.13% |
|  | 5HT_2_ | Serotonergic | 0.06% |
|  | D_1_ x GM | Dopaminergic | 0.10% |
|  | D_1_ x MD | Dopaminergic | 0.07% |
|  | D_1_ | Dopaminergic | 0.06% |
|  | offset | Non-Receptor | 0.06% |
|  | t1/t2 | Non-Receptor | 0.04% |
| fALFF | GABA_B_ x SPECT | GABAergic | 0.06% |
|  | M_3_ x GM | Cholinergic | 0.06% |
|  | α_4_β_2_ x t1/t2 | Cholinergic | 0.05% |
|  | 5HT_2_ x MD | Serotonergic | 0.08% |
|  | 5HT_1A_ x t1/t2 | Serotonergic | 0.03% |
|  | GM | Non-Receptor | 0.04% |
|  | FA | Non-Receptor | 0.06% |
|  | t1/t2 | Non-Receptor | 0.04% |
| SPECT | AMPA | Glutamatergic | 0.04% |
|  | NMDA | Glutamatergic | 0.08% |
|  | Kainate | Glutamatergic | 0.12% |
|  | GABA_B_ x GM | GABAergic | 0.05% |
|  | Bz site x fALLF | GABAergic | 0.05% |
|  | GABA_B_ x FA | GABAergic | 0.07% |
|  | α_4_β_2_ x SPECT | Cholinergic | 0.14% |
|  | α_1_ x SPECT | Adrenergic | 0.04% |
|  | α_2_ x SPECT | Adrenergic | 0.07% |
|  | α_2_ | Adrenergic | 0.09% |
|  | 5HT_2_ x fALLF | Serotonergic | 0.09% |
|  | D_1_ x FA | Dopaminergic | 0.03% |
| FA | Kainate x GM | Glutamatergic | 0.10% |
|  | NMDA x SPECT | Glutamatergic | 0.10% |
|  | Kainate x FA | Glutamatergic | 0.09% |
|  | Kainate | Glutamatergic | 0.12% |
|  | GABA_A_ x SPECT | GABAergic | 0.04% |
|  | GABA_B_ | GABAergic | 0.07% |
|  | α_1_ x GM | Adrenergic | 0.06% |
|  | α_2_ x fALLF | Adrenergic | 0.05% |
|  | α_1_ x SPECT | Adrenergic | 0.09% |
|  | α_1_ x t1/t2 | Adrenergic | 0.06% |
|  | 5HT_1A_ | Serotonergic | 0.08% |
|  | D_1_ x MD | Dopaminergic | 0.07% |
| MD | Kainate x MD | Glutamatergic | 0.04% |
|  | Bz site x fALLF | GABAergic | 0.06% |
|  | Bz site x SPECT | GABAergic | 0.07% |
|  | GABA_A_ x FA | GABAergic | 0.07% |
|  | Bz site x FA | GABAergic | 0.04% |
|  | GABA_B_ x MD | GABAergic | 0.16% |
|  | Bz site | GABAergic | 0.06% |
|  | GABA_B_ | GABAergic | 0.06% |
|  | M_2_ x fALLF | Cholinergic | 0.05% |
|  | M_1_ x SPECT | Cholinergic | 0.03% |
|  | M_2_ x FA | Cholinergic | 0.07% |
|  | M_1_ x t1/t2 | Cholinergic | 0.06% |
|  | M_2_ x t1/t2 | Cholinergic | 0.06% |
|  | M_2_ | Cholinergic | 0.06% |
|  | M_3_ | Cholinergic | 0.09% |
|  | α_2_ x fALLF | Adrenergic | 0.04% |
|  | 5HT_1A_ x GM | Serotonergic | 0.05% |
|  | 5HT_1A_ x t1/t2 | Serotonergic | 0.04% |
|  | 5HT_2_ | Serotonergic | 0.06% |
|  | D_1_ x SPECT | Dopaminergic | 0.07% |
|  | t1/t2 | Non-Receptor | 0.06% |
| t1/t2 | AMPA x SPECT | Glutamatergic | 0.07% |
|  | NMDA x FA | Glutamatergic | 0.07% |
|  | GABA_A_ x GM | GABAergic | 0.04% |
|  | GABA_B_ x GM | GABAergic | 0.04% |
|  | Bz site x SPECT | GABAergic | 0.06% |
|  | GABA_B_ x SPECT | GABAergic | 0.05% |
|  | M_1_ x GM | Cholinergic | 0.08% |
|  | M_3_ x GM | Cholinergic | 0.14% |
|  | M_2_ x SPECT | Cholinergic | 0.04% |
|  | α_4_β_2_ x MD | Cholinergic | 0.06% |
|  | M_1_ | Cholinergic | 0.16% |
|  | M_3_ | Cholinergic | 0.15% |
|  | α_1_ x GM | Adrenergic | 0.03% |
|  | α_1_ x SPECT | Adrenergic | 0.07% |
|  | α_1_ x FA | Adrenergic | 0.04% |
|  | α_1_ x MD | Adrenergic | 0.08% |
|  | 5HT_2_ x GM | Serotonergic | 0.07% |
|  | 5HT_2_ x MD | Serotonergic | 0.05% |
|  | D_1_ x MD | Dopaminergic | 0.06% |
|  | D_1_ x t1/t2 | Dopaminergic | 0.07% |
|  | GM | Non-Receptor | 0.06% |

**Supplementary Table S7:** *Total MCM parameter-clinical co-variance explained by receptor type in PD patients (via each SVD component).*

| **Receptor Type** | **Total Variance Explained in PC1** | **Total Variance Explained in PC2** |
| --- | --- | --- |
| **Glutamatergic** | 4.85% | 1.19% |
| **GABAergic** | 5.97% | 1.24% |
| **Cholinergic** | 4.77% | 1.74% |
| **Adrenergic** | 2.02% | 0.88% |
| **Serotonergic** | 3.77% | 0.75% |
| **Dopaminergic** | 1.16% | 0.53% |

**Supplementary Table S8:** *Performance gain due to the inclusion of receptor maps, and the p-value from a two-sample t-test for each modality.*

| **Imaging Modality** | **Average Gain in R^2^** | **P-value** |
| --- | --- | --- |
| **GM** | 35.6% ± 10.7% | P<10^-26^ |
| **fALFF** | 18.8% ± 8.0% | P<10^-12^ |
| **SPECT** | 20.1% ± 12.3% | P<10^-8^ |
| **FA** | 21.7% ± 11.8% | P<10^-10^ |
| **MD** | 19.0% ± 9.0% | P<10^-8^ |
| **t1/t2** | 17.1% ± 9.3% | P<10^-8^ |

**Supplementary Table S9:** *Performance gain due to true receptor distributions over null maps, and p-value of the true receptor data model belonging to the null distribution.*

| **Imaging Modality** | **Average Gain in R^2^** | **P-value** |
| --- | --- | --- |
| **GM** | 13.4% ± 5.3% | P<0.001 |
| **fALFF** | 7.3% ± 4.5% | P<0.001 |
| **SPECT** | 6.7% ± 4.3% | P<0.001 |
| **FA** | 7.5% ± 5.3% | P<0.001 |
| **MD** | 5.3% ± 4.0% | P<0.001 |
| **t1/t2** | 6.0% ± 3.5% | P<0.001 |

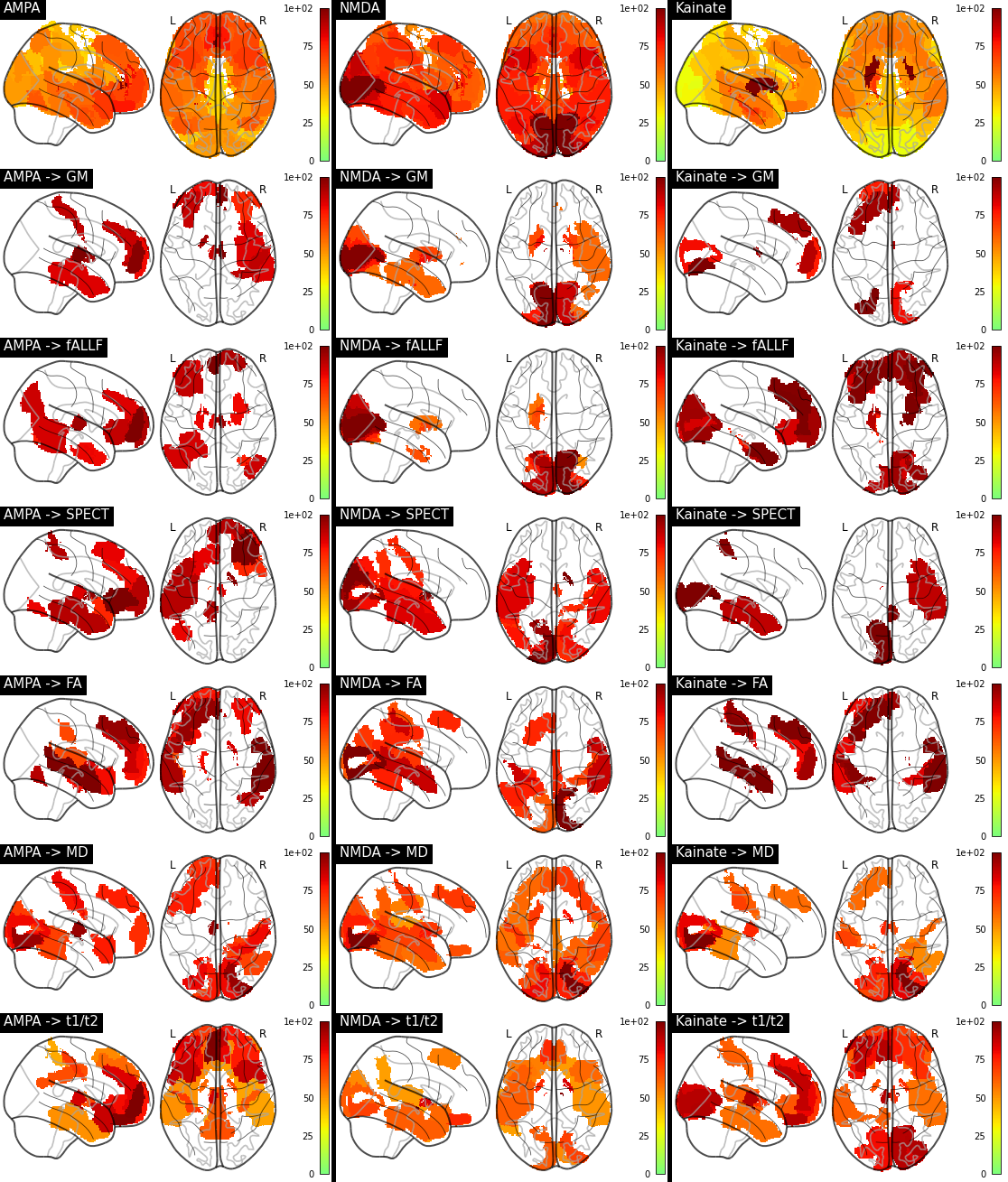
**Supplementary Figure S1:** *Glutamatergic receptor influence maps. The first row contains the density maps, re-scaled for visualization. Influence maps are re-scaled to arbitrary units for visualization, and show only regions with significant z-scores (P<0.05) of Wilcoxon rank-sum statistics relative to the Wilcoxon statistics due to null distributions.*

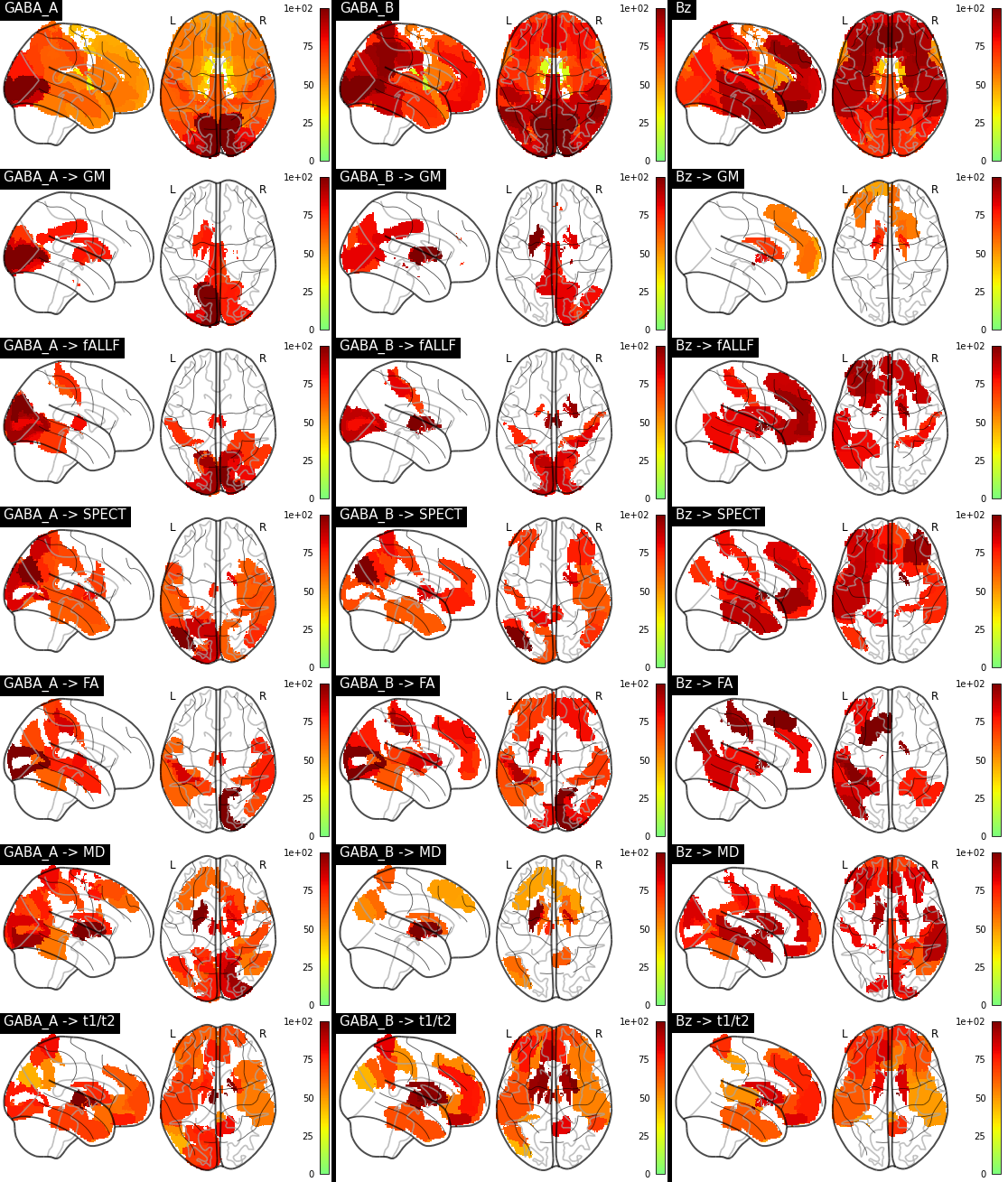

**Supplementary Figure S2:** *GABAergic receptor influence maps*

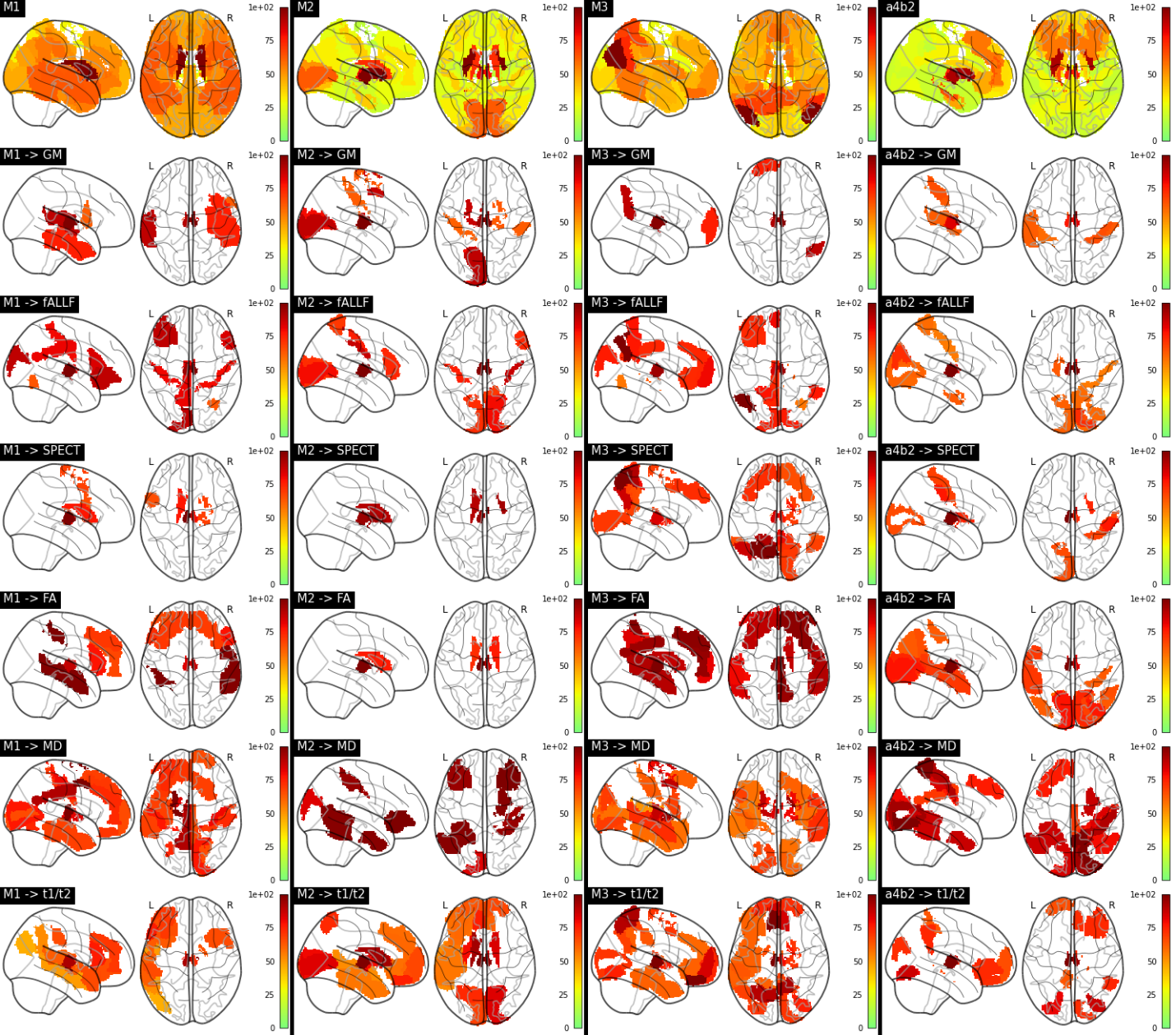

**Supplementary Figure S3:** *Cholinergic receptor influence maps*

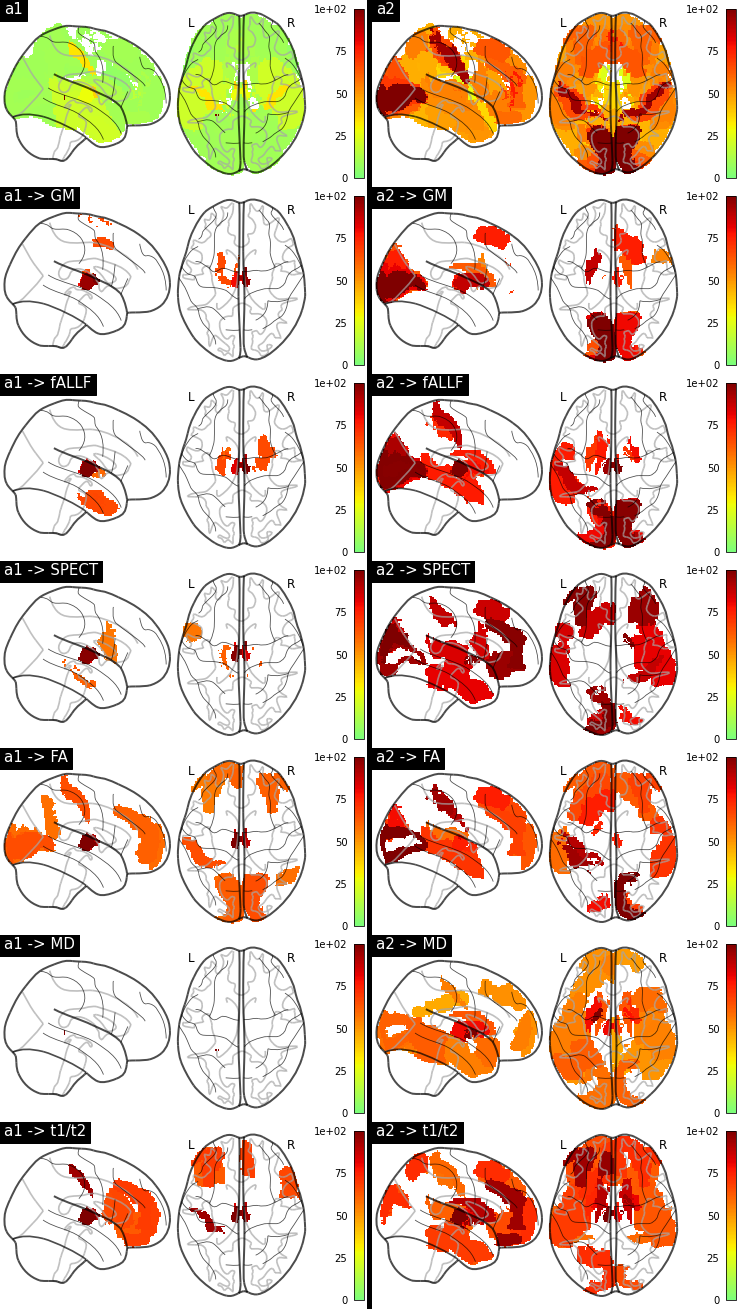

**Supplementary Figure S4:** *Adrenergic receptor influence maps*

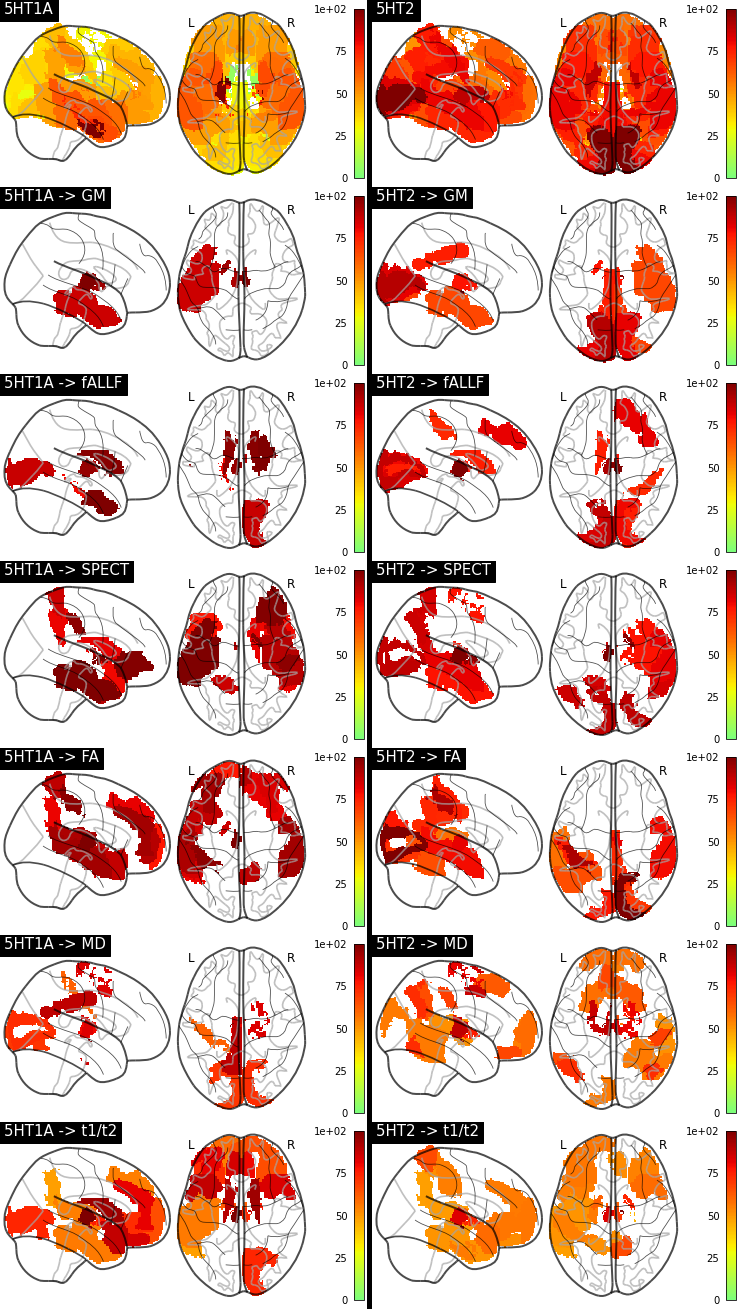

**Supplementary Figure S5:** *Serotonergic receptor influence maps*

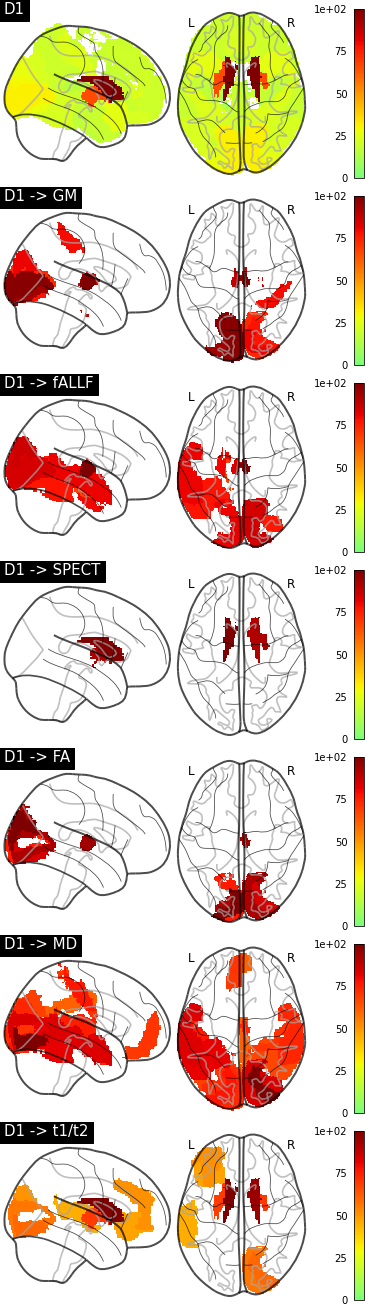

**Supplementary Figure S6:** *Dopaminergic receptor influence maps*
